# Simple and Versatile Pharmacokinetic Model for Radioligand Therapy

**DOI:** 10.1101/2024.06.13.24308814

**Authors:** Adrian Jun Zounek, Alexis Nikolai Zounek

## Abstract

In recent years radioligand therapy has emerged as an effective treatment modality for various solid malignancies, with pharmacokinetic modeling being routinely used for absorbed dose calculation and patient-specific therapy planning. Exemplary time-activity curves of FAP-targeted radioligands in a mouse model are accurately fitted by a sum of right skew biexponential distributions with four adjustable parameters in total. This type of modeling function is versatile and also suitable for conventional drugs. For further insight, an auxiliary equation is derived that relates tumor clearance to FAP expression and the radioligand dissociation constant.

---

Radioligand therapy (RLT) has been recognized as a promising treatment modality for solid tumors as demonstrated by the March 2022 FDA approval of [^177^Lu]-PSMA-617 (Pluvicto®, Novartis AG, Basel, Switzerland) for the treatment of metastatic castration-resistant prostate cancer (mCRPC). Worldwide efforts are underway to develop new radioligands and expand the scope of RLT. In order to support these efforts and to gain a better understanding of RLT, various physiologically based pharmacokinetic (PBPK) models have been devised and adopted for patient-specific dosimetry and therapy planning. Despite their high utility PBPK models often include a large number of adjustable parameters with limited interpretability.

In this article, a simple and versatile PBPK model with only four adjustable parameters is presented. The model requires a blood time-activity curve (TAC), which can be obtained based on image data or using automatic or manual blood sampling. The two main parameters of the PBPK model refer to two exponential rate constants, which are similar to the eigenvalues of a multi-compartment model.

An N-compartment model includes 2N-1 micro or tissue rate constants and has N eigenvalues and N amplitudes for each compartment (N^2^ amplitudes total). The model eigenvalues and amplitudes of each compartment may be determined from the corresponding time-activity or time-concentration curves via nonlinear regression. However, this approach often proves to be problematic due to overfitting and leads to inaccurate results - especially for small eigenvalues, i.e. slow processes.

The proposed PBPK model uses a simple and numerically stable regression method to determine the eigenvalues and amplitudes based on the TACs of the blood and tumor compartment. The TAC of the tumor compartment is fitted with functions of type 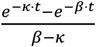, which corresponds to a right skewed biexponential distribution [1].

For therapeutic use, the primary goal is to describe the TAC of the tumor compartment with high reliability. Knowledge of tissue rate constants is of great interest for basic understanding but is not clinically required.

The article further presents a formula that describes radioligand clearance from tumor tissue by a simple function of affinity, target receptor concentration and “regular” pharmacokinetic clearance without receptor interference. To the best of our knowledge, this is the first time such a relationship has been derived.

RLT uses radioligands that preferentially accumulate in tumor tissue and expose them to an elevated radiation dose. Radioligand compounds comprise a targeting moiety or ligand coupled to a chelator via a spacer. The ligand binds to a biological receptor, such as prostate specific membrane antigen (PSMA), fibroblast activation protein (FAP) or a somatostatin receptor (SSTR) that is overexpressed in neoplastic lesions. The ligand can be a monoclonal antibody, a peptide or a small peptidomimetic molecule. The chelator is suited for complexation of a diagnostic or therapeutic radioisotope, such as ^68^Ga, ^177^Lu, ^90^Y, ^225^Ac having a half-life of 68 min, 6.65 d, 64.1 h and 9.92 d, respectively. Depending on the radioisotope the ionizing radiation dose is delivered primarily by high-energy photons, electrons or alpha-particles. The spacer couples the chelator and therewith complexed radioisotope to the targeting ligand while at the same time mitigating the interference of the chelator with ligand binding to its target receptor.

RLT uses radioligands with rapid systemic excretion and, conversely, high uptake and long retention in tumor tissue. For the latter, the affinity of the radioligand to its target receptor plays a crucial role. Affinity is quantified by its reciprocal, the dissociation constant *K*_*D*_, preferably in units of nM (10^-9^ mol·L^-1^). For the clinically approved radiotracer [^177^Lu]-PSMA-617, Wen et al. [2] report values of *K*_*D*_ = 4.7 nM and k_off_ = 3.44 × 10^-4^ s^-1^. k_off_ denotes the equilibrium dissociation rate, which in the case of [^177^Lu]-PSMA-617 corresponds to a binding half-life of ln(2)/3.44 × 10^4^ s = 33.6 min. The latter is 285 times smaller than the 9751.0 min (6.65 d) half-life of the therapeutic radioisotope ^177^Lu. The large difference between the binding half-life of radioligands and therapeutic radioisotopes used for them is remarkable and deserves explanation. Due to 90 % cellular internalization [^177^Lu]-PSMA-617 has a retention half-life in tumor tissue in the range from 68,0 h to 74,5 h which exceeds its binding half-life by a factor of about 127 (cf. Galler et al. [3], Fig. 8 legend). However, for radioligands that bind to FAP-positive CAFs, the internalization ratio appears to be considerably lower [4, 5]. Accordingly, the retention time in tumor tissue is largely limited by radioligand dissociation from extracellular FAP.

Without cellular internalization, retention in tumor tissue depends on radioligand pharmacokinetics and *K*_*D*_ or affinity to the target receptor R. As shown in appendix A, the rate constant *κ*_*e*_ for radioligand clearance from tumor tissue is linked to the dissociation constant *K*_*D*_ and the intratumoral receptor concentration [*R*]_*t*_ via the following relationship:

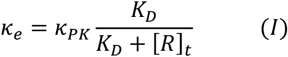

wherein *κ*_*PK*_ denotes the physiological clearance rate in the absence of the target receptor R. According to equation (I) *κ*_*e*_ tends towards *κ*_*PK*_ for large dissociation constant *K*_*D*_ ≫ [*R*]_*t*_ i.e. low affinity. If *κ*_*PK*_, *K*_*D*_ and [*R*]_*t*_ are known, equation (I) can be used to calculate *κ*_*e*_. Conversely, the intratumoral receptor concentration [*R*]_*t*_ may be determined based on *κ*_*e*_, *κ*_*PK*_ and *K*_*D*_.

The physiological clearance rate *κ*_*PK*_ in tumor tissue largely depends on intercellular diffusion and convection. Convection can be accelerated due to fenestrated vasculature or shunts between arterial and venous capillaries, as commonly found in tumor tissue. In contrast to normal tissue, lymphatic clearance in tumor tissue is severely impaired. Regardless of the different physiology of tumor and normal tissue, the latter can serve as a useful approximation for *κ*_*PK*_. Alternatively, *κ*_*PK*_ can be experimentally determined using multilayered cell cultures or multicellular spheroids.

As can be gathered from equation (I) the effective clearance rate *κ*_*e*_ can be lowered by reducing either *κ*_*PK*_ or *K*_*D*_. Modification of radioligand chemistry affects both *κ*_*PK*_ and *K*_*D*_. Therefore, it may be difficult to distinguish or decouple the effect of reduced *K*_*D*_ from the effect of altered *κ*_*PK*_ in tumor tissue and pharmacokinetics in healthy tissue. This interdependence must be heeded especially if clearance from other tissues such as liver or bone marrow is slow.

Reducing *κ*_*PK*_ is likely to prolong radioligand retention in healthy tissue, which is generally undesirable. Notwithstanding, strategies aimed at prolonging retention in the blood compartment can improve efficacy, as recently shown using radioligands equipped with an additional albumin binding moiety [6-9].

The proposed PBPK model is based on the familiar differential equation (II)

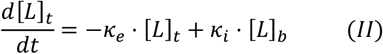

comprising a source term *κ*_*i*_ ⋅ [*L*]_*b*_ and a clearance term −*κ*_*e*_ ⋅ [*L*]_*t*_ wherein the source term is the product of the radioligand uptake rate *κ*_*i*_ times the concentration [*L*]_*b*_ in blood. Clearance is likewise described by the product of the clearance rate *κ*_*e*_ times the intratumoral concentration [*L*]_*t*_. Integration of equation (II) yields the general solution:

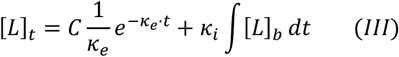

If [*L*]_*b*_ can be represented by a single exponential function of type

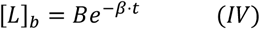

the solution of equation (III) takes the form:

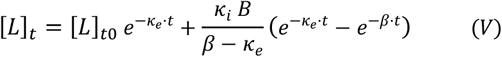

wherein the initial tumor ligand activity [*L*]_*t*0_ is equal to zero in most cases and the term 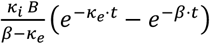 corresponds to a right skewed biexponential distribution [1].

The solution according to equation (V) tacitly ignores radioligand clearance from tumor tissue into the venous blood compartment and may violate mass conservation. However, this neglect is justified for clinical patients in whom tumor lesions typically constitute a small portion of body mass. Furthermore, as the beneath examples show, a model with two terms of type 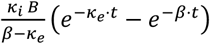 and four adjustable parameters can describe tumor TACs in mouse xenografts despite abnormally large tumor-to-body mass ratios.

Taking mass conservation into account, on the other hand, requires a multi-compartment model with four or more compartments and numerical solution of a corresponding system of coupled differential equations with seven or more adjustable parameters. Apart from notoriously unstable numerical solutions and overfitting, an increase in the number of adjustable parameters makes meaningful interpretation difficult.

To demonstrate the use and versatility of the proposed PBPK model, two terms of the type 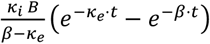 were used to fit experimental TACs measured by Wang et al. in mouse xenograft models for four different FAP radioligands [10]. In addition to the well-known reference compound FAPI-04, Wang et al. report experimental TACs for three newly designed radioligands denoted as “Ga-3-3”, “Ga-6-3” and “Ga-8-1”. For a detailed description of the latter the reader is referred to their article.

Growing evidence supports the notion that FAP-expressing cancer associated fibroblasts (CAF^FAP+^) suppress anti-cancer immune response and promote tumor progression and metastasis [11]. Accordingly, FAP constitutes a promising target receptor for chemotherapy and RLT. Recent clinical results indicate that FAP-targeted RLT can be an effective and benign treatment modality [12-14].

The systematic study by Wang et al. can provide valuable clues for improving FAP-targeted radioligands. Their experimental data highlight the quite variable effects of radioligand pharmacokinetics on TACs and tumor dose, making validation of a PBPK model challenging. For these reasons, their data was chosen to test the proposed PBPK model.

First the blood and tumor TACs of Fig. 5b of Wang et al. for radioligands FAPI-04, Ga-3-3, Ga-6-3 and Ga-8-1 were digitized. Each of the four digitized blood TACs was then fitted by a triexponential function of type:

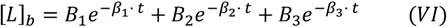

The fitted model curves and corresponding parameter values are presented in Fig. 1 and Table 1. Integration of equation (VI) yields the area under the curve for the blood concentration [*L*]_*b*_ as

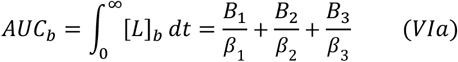

**Table 1:**
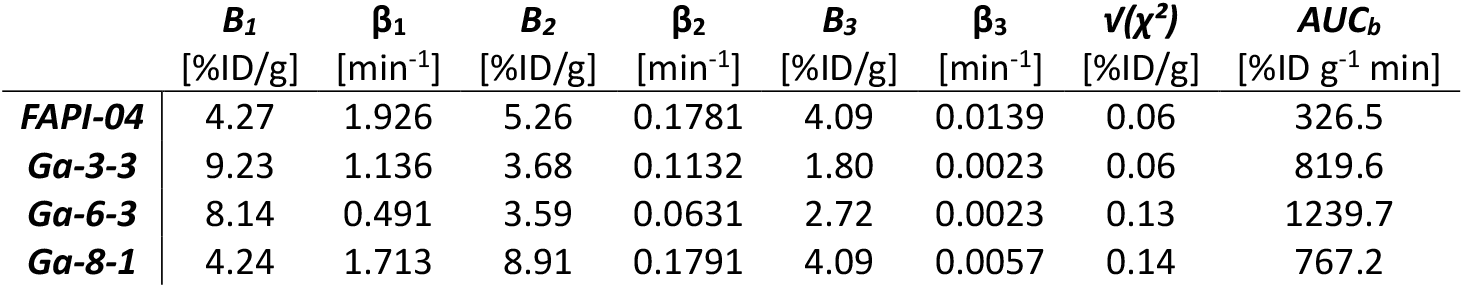
Fit parameters of the blood TACs.

**Fig. 1:**
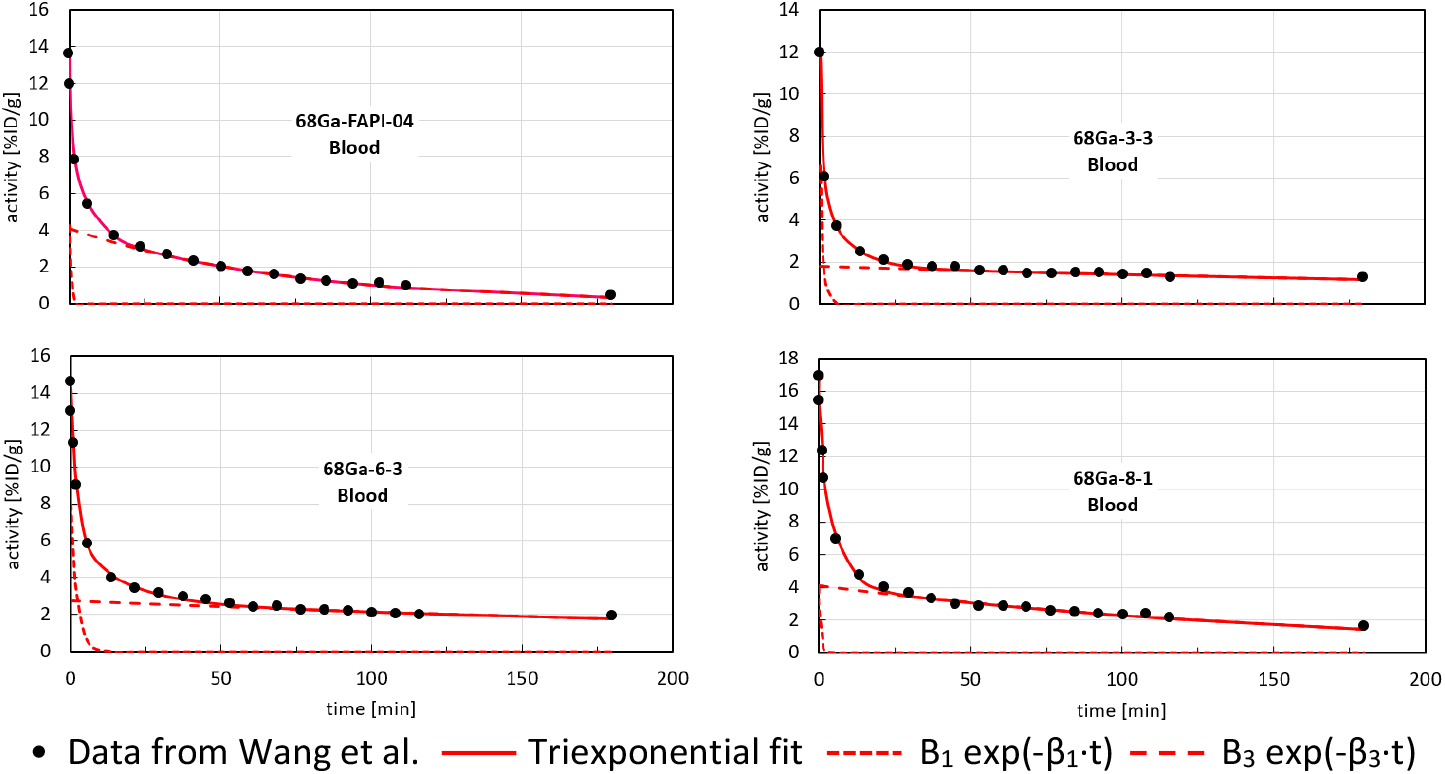
Blood TACs fitted to the data measured by Wang et al. [10].

The function used to fit the tumor TACs is represented by the following formula (VII):

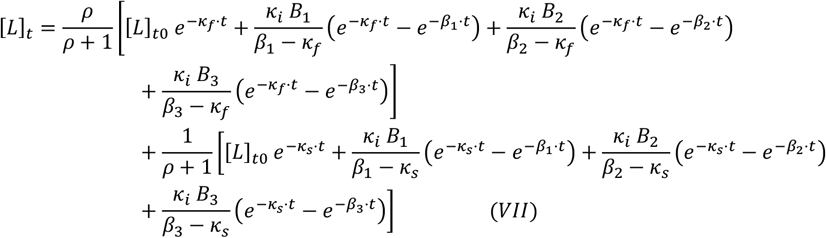

comprising exponential terms according to equation (V) and four adjustable parameters *κ*_*i*_, *κ*_*f*_, *κ*_*s*_, *ρ*. The exponential rate constants *κ*_*f*_, *κ*_*s*_, *β*_1_, *β*_2_, *β*_3_ are similar to the eigenvalues of a 5-compartment model and do not directly correspond to physiological tissue rate constants, but rather are complicated functions thereof. As can be easily shown through differentiation, the intratumoral radioligand activity [*L*]_*t*_ of equation (VII) satisfies differential equation (II). Integration of equation (VII) yields the area under the curve for the tumor concentration [*L*]_*t*_ as

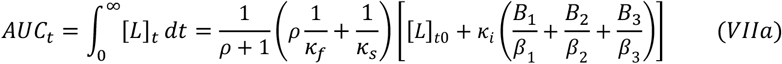

For simplicity, in the following, the parameters in equations (VI), (VIa), (VII), (VIIa) are assumed to comprise a general scaling factor and to be given in units of

‒ [%ID/g] for [*L*]_*t*_, [*L*]_*t*0_, [*L*]_*b*_, *B*_1_, *B*_2_, *B*_3_; and

‒ [%ID g^-1^ min] for *AUC*_*b*_, *AUC*_*t*_.

This assumption has no influence on key pharmacokinetic parameters, such as the rate constants *κ*_*i*_, *κ*_*f*_, *κ*_*s*_ and the dimensionless partition factor *ρ*. In addition, the use of a general scaling factor is common practice when dealing with TACs.

Equation (VII) formally corresponds to a sum of five exponential terms. If three exponential rate constants can be determined based on a blood TAC, the number of nominally adjustable parameters drops from ten to seven. The proposed PBPK model further reduces this number to four adjustable parameters. In a clinical setting the mass of the tumor compartment is small compared to healthy tissue. Accordingly, clearance from the tumor compartment has minor impact on the blood compartment. Under these “normal” conditions the blood compartment mainly exhibits renal and hepatobiliary excretion and to some extent radioligand recirculation due to clearance from healthy tissue. Contrary thereto, the blood TACs of Wang et al. are noticeably influenced by clearance from the abnormally large xenograft tumors. As consequence the triexponential fit to the blood TACs of Wang et al. involves at least one exponential term that mirrors tumor clearance.

Based on equation (VII) each of the four digitized tumor TACs of Wang et al. was fitted. The results are shown in Fig. 2 and Table 2. The fitted model curves generally agree well with the experimental data of Wang et al. as evidenced by the χ^2^ values reported in Table 2. This suggests that the proposed PBPK model provides a fairly accurate description of experimental tumor TACs and can be used to extrapolate to times that exceed the measurement time of 180 minutes. However, this assumption needs to be confirmed by experiments over longer periods of time.

**Table 2:** Fit parameters of the tumor TACs.

| | $\rho$ | $\kappa_i$<br>[min<sup>-1</sup>] | $\kappa_f$<br>[min<sup>-1</sup>] | $\kappa_s$<br>[min<sup>-1</sup>] | $v(\chi^2)$<br>[%ID/g] | $AUC_t$<br>[%ID g<sup>-1</sup> min] | $AUC_t / AUC_b$ |
| --- | --- | --- | --- | --- | --- | --- | --- |
| **FAPi-04** | 10.30 | 0.30443 | 0.14298 | 0.00220 | 0.40 | 4859.3 | 14.9 |
| **Ga-3-3** | 1.19 | 0.45408 | 0.17705 | 0.01907 | 0.27 | 10188.4 | 12.4 |
| **Ga-6-3** | 2.87 | 0.27041 | 0.15377 | 0.00501 | 0.60 | 19204.3 | 15.5 |
| **Ga-8-1** | 3.45 | 0.16710 | 0.05505 | 0.00215 | 0.42 | 15767.2 | 20.6 |

**Fig. 2:**
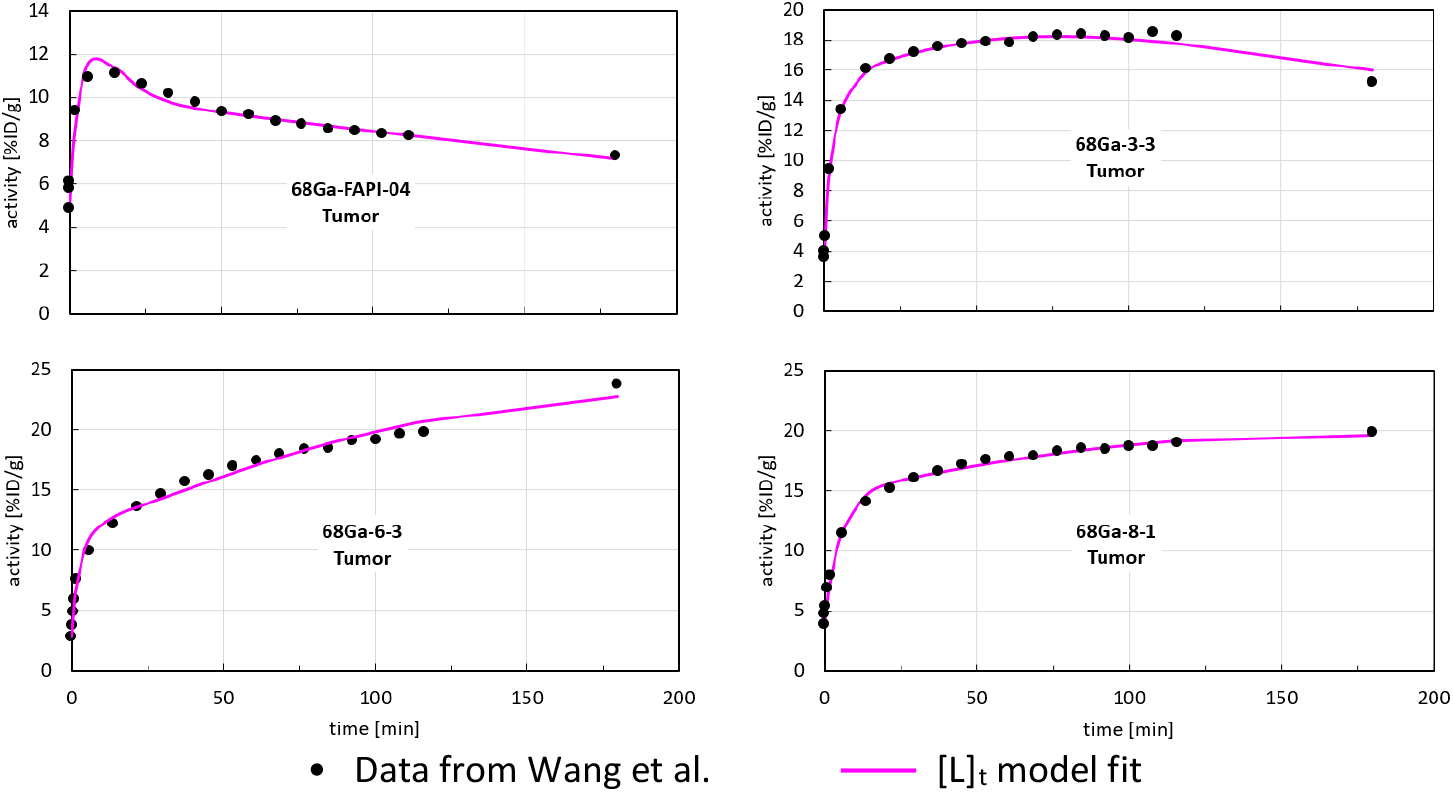
Tumor TACs fitted to the data measured by Wang et al. [10].

Despite the limitations mentioned above, the proposed PBPK model affords robust determination of model parameters *κ*_*i*_, *κ*_*f*_, *κ*_*s*_, *ρ* and thereon based calculation of tumor dose (*AUC*_*t*_) and tumor-to-blood dose ratio (*AUC*_*t*_/*AUC*_*b*_). It appears that the robustness of the proposed PBPK model is owed to the small number of adjustable parameters and that *κ*_*i*_ and *ρ* largely correspond to scaling factors with limited influence on the shape of the fitted TACs.

The proposed PBPK model can be advantageously used in the clinic, especially in settings where blood TACs are recorded simultaneously with dynamic PET, and may contribute to the understanding of pharmacokinetics and its impact on radioligand effectiveness.

As shown in Appendix B, a 5-compartment model cannot adequately describe the experimental results of Wang et al. This finding calls for caution when using a multi-compartment model and urges thorough analysis and comparison of the TACs and rate constants of all relevant tissues, with particular attention to liver and bone marrow. With this limitation in mind, the exponential rate constants derived by the proposed PBPK model are assumed to behave similarly to the eigenvalues *λ*_1≤*j*≤5_ of a 5-compartment model and represent bounds on the tissue rate constants.

The eigenvalues *λ*_1≤*j*≤*N*_ of a general N-compartment model are complicated functions of the underlying tissue uptake and clearance rate constants *k*_1≤*i*≤2*N*−1_. For given tissue rate constants *k*_1≤*i*≤2*N*−1_, the eigenvalues *λ*_1≤*j*≤*N*_ satisfy the condition

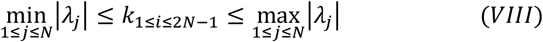

FAP radioligands are excreted from the central blood compartment via the renal or hepatobiliary pathway either directly or after tissue perfusion and reentry, i.e. clearance into the blood compartment. Accordingly, the blood and tumor TACs are complex functions of tissue clearance and systemic excretion.

Hydrophilic radioligands tend to have high tissue diffusivity and are preferably excreted via the kidneys. Conversely, lipophilic radioligands perfuse slowly and are more likely to be excreted via the hepatobiliary pathway.

Heuristically, blood and tissue TACs are expected to comprise a ligand or activity component that has traveled a maximal path between injection and systemic excretion. However, depending on the amount of this component it may not noticeably contribute to either the blood or tumor TAC.

Based on this reasoning, the larger of the exponential rate constants *β*_3_ and *κ*_*s*_ reported in Table 1 and Table 2 is considered as approximation 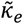 for the effective tumor clearance rate *κ*_*e*_, i.e. 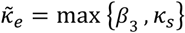. If *κ*_*PK*_ and *K*_*D*_ are known, 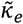 can be substituted into equation (I) to estimate the intratumoral receptor concentration [*R*]_*t*_ according to the formula

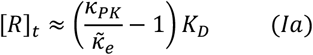

Table 3 lists logP, IC_50_, 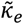 and corresponding binding half-lives 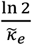 for radioligands FAPI-04, Ga-3-3, Ga-6-3, Ga-8-1.

**Table 3:** Radioligand pharmacokinetic parameters (*logP for FAPI-04 adopted from Yang et al., page 2334, right column, 1st paragraph [4]).

| **Radioligand** | **$\log P^*$** | **$IC_{50} [nM]$** | **$\tilde{\kappa}_e [min^{-1}]$** | **$\frac{\ln 2}{\tilde{\kappa}_e} [min]$** |
| --- | --- | --- | --- | --- |
| **Ga-3-3** | -3.23 | 2.2 | 0.019071 | 36.3 |
| **FAPI-04** | -2.42 | 3.8 | 0.013870 | 50.0 |
| **Ga-8-1** | -2.55 | 3.3 | 0.005725 | 121.1 |
| **Ga-6-3** | -1.51 | 1.9 | 0.005007 | 138.4 |

The above data does not exhibit a systematic correlation between logP or IC_50_ and the binding half-life 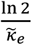. Obviously, though, the naphthol moiety (cf. Fig. 3) incorporated in Ga-6-3 and Ga-8-1 substantially prolongs the binding half-life 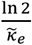 relative to FAPI-04 and Ga-3-3. It appears that the prolongation imparted by naphthol is attributable to increased lipophilicity and protein affinity. Surprisingly, the latter does not seem to be FAP specific.

**Fig. 3:**
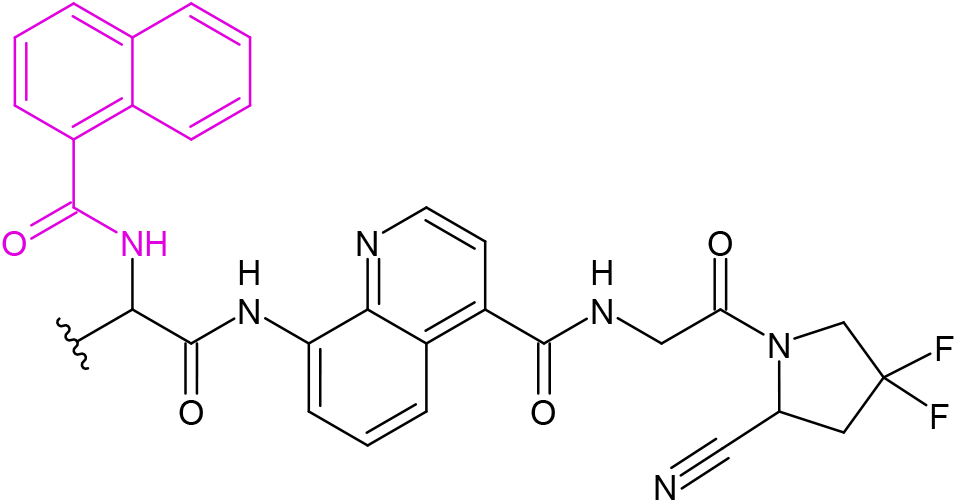
Naphthol moiety (marked in magenta) incorporated in radioligands Ga-6-3 and Ga-8-1 [10].

The rather high IC_50_ value of Ga-8-1 and the strong uptake of Ga-6-3 in liver tissue point to a nonspecific pharmacokinetic mechanism.

As discussed in Appendix B, the authors attempted to reproduce the tumor TACs of Wang et al. based on a 5-compartment model with nine adjustable rate constants. For radioligands Ga-3-3, Ga-6-3 and Ga-8-1 it was not possible to fit the experimental data in a satisfactory manner. It appears, that despite large degrees of freedom, a 5-compartment model cannot adequately describe the particular pharmacokinetics of these radioligands.

## Data Availability

All data produced in the present study are available upon reasonable request to the authors

## Appendix A

In thermal equilibrium, the same number of complexes RL are formed and dissociate in every unit of time and volume. The number of newly formed and dissociating complexes per unit of time and volume is proportional to the receptor concentration [*R*] and the ligand concentration [*L*] with the proportionality constant k_on_ and proportional to the complex concentration [*RL*] with the proportionality constant k_off_, respectively.

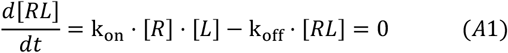

Accordingly, we get

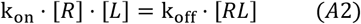

and

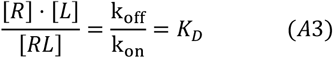

with the dissociation constant *K*_*D*_ (or reciprocal of the affinity). Transforming equation (A3) yields:

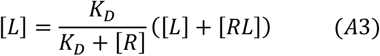

Radioligand clearance from tumor tissue is proportional to the concentration [*L*] of unbound radioligand and the “regular” pharmacokinetic clearance rate *k*_*PK*_ without receptor interference

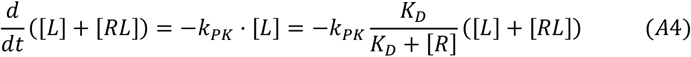

This gives us the effective clearance rate

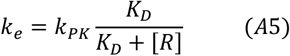

According to equation (A5), *k*_*e*_ < *k*_*PK*_ for all values of *K*_*D*_ and [*R*]. For low affinity, i.e. *K*_*D*_ ≫ [*R*], *k*_*e*_ tends toward *k*_*PK*_ (*k*_*e*_ → *k*_*PK*_). Conversely, for very high affinity *K*_*D*_ → 0, *k*_*e*_ tends toward 0 (*k*_*e*_ → 0).

A typical scenario with *K*_*D*_ = 4.7 nM and [*R*] = 16 nM yields 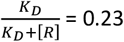, i.e. the effective tumor clearance rate *k*_*e*_ is smaller (slower) by a factor of 4.4 relative to the “regular” pharmacokinetic clearance rate *k*_*PK*_. For fixed *K*_*D*_ and [*R*] the effective clearance rate *k*_*e*_ can be altered by changing the pharmacokinetic clearance rate *k*_*PK*_. Generally, a modification of radioligand chemistry affects both *K*_*D*_ and *k*_*PK*_.

For therapeutically effective radioligand retention in tumor tissue a 1:10 ratio according to the formula

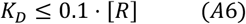

can serve as rule of thumb.

## Appendix B

PBPK models for RLT are usually based on a multi-compartment model and respective system of coupled linear differential equations.

The PBPK model proposed in this article is based on equation (VII) which formally corresponds to a sum of five exponential functions. Hence, the authors sought to construct an analogous 5-compartment model.

For this purpose, three different 5-compartment models with characteristic matrices 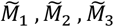 were examined.

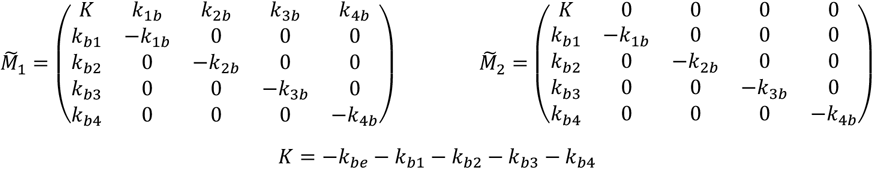

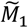 and 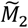 represent models with one blood and four tissue compartments with corresponding rate constants *k*_*be*_, *k*_1*b*_, *k*_2*b*_, *k*_3*b*_, *k*_4*b*_ for blood excretion and tissue clearance, and *k*_*b*1_, *k*_*b*2_, *k*_*b*3_, *k*_*b*4_ for tissue uptake. 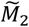 differs from 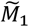 in that recirculation from the four tissue compartments into the blood compartment is neglected.

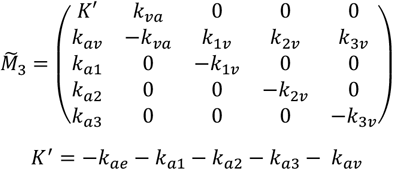

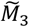 corresponds to a model with two compartments for arterial and venous blood and three tissue compartments. Therein *k*_*ae*_, *k*_*av*_, *k*_*va*_ are the rate constants for arterial excretion and arterial-venous recirculation, whereas *k*_*a*1_, *k*_*a*2_, *k*_*a*3_, *k*_1*v*_, *k*_2*v*_, *k*_3*v*_ denote rates for tissue uptake from arterial blood and clearance into venous blood, respectively.

The rate constants or coefficients of each of matrices 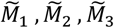 were fitted to the tumor TAC of each of radioligands FAPI-04, Ga-3-3, Ga-6-3 and Ga-8-1 using a Nelder-Mead optimization algorithm (https://en.wikipedia.org/wiki/Nelder-Mead_method). The corresponding blood TACs were not included in order to alleviate the optimization restraints.

The rate constants or coefficients of each of matrices 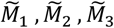 are equivalent to the rate constants of a system of five coupled differential equations. Rather than using numerical integration based on a Runge-Kutta method [15], the five eigenvalues and eigenvectors {*λ*_*j*_, 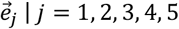} of the matrices 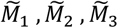 were numerically determined in each iteration of the optimization procedure. A linear superposition according to formula (IB)

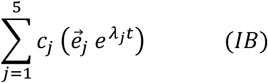

with five coefficients {*c*_*j*_ | *j* = 1, 2, 3, 4, 5} constitutes a solution to the system of coupled differential equations. The coefficients *c*_*j*_ are determined by solving the linear system of equations for the boundary conditions at time t = 0.

The eigenvalue approach avoids the instabilities that frequently plague the numerical integration of “stiff” differential equation systems with rate constants that differ by two or three orders of magnitude.

For the first model with characteristic matrix 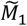 the results of the optimization procedure are presented in Table B 1 and Fig. B 1. While it was possible to match the tumor TACs of FAPI-04 and Ga-8-1 with satisfactory conformance the agreement for Ga-3-3 and Ga-6-3 is poor, despite exclusion of the blood TACs and the rather large degrees of freedom. Moreover, for Ga-3-3, Ga-6-3 and Ga-8-1 the resultant rate constant for tumor clearance tends towards zero, i.e. infinite retention, which obviously contradicts experimental results obtained by Wang et al. and others.

**Table B 1:**
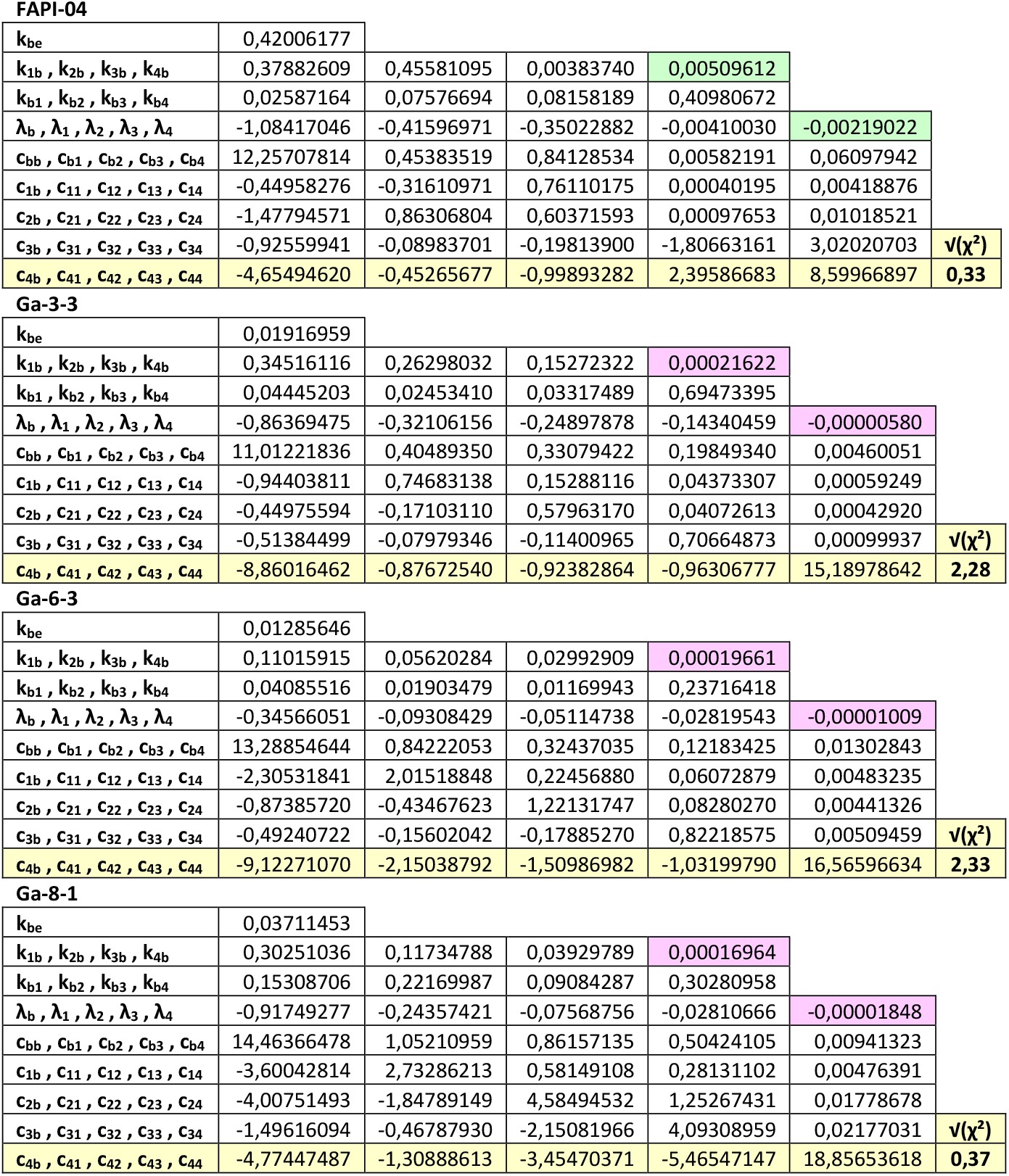
Coefficients of the optimized 5-compartment model.

**FAPI-04**
|  |  |  |  |  |  |
| --- | --- | --- | --- | --- | --- |
| $k_{be}$ | 0,42006177 | | | | |
| $k_{1b}, k_{2b}, k_{3b}, k_{4b}$ | 0,37882609 | 0,45581095 | 0,00383740 | 0,00509612 | |
| $k_{b1}, k_{b2}, k_{b3}, k_{b4}$ | 0,02587164 | 0,07576694 | 0,08158189 | 0,40980672 | |
| $\lambda_b, \lambda_1, \lambda_2, \lambda_3, \lambda_4$ | -1,08417046 | -0,41596971 | -0,35022882 | -0,00410030 | -0,00219022 |
| $c_{bb}, c_{b1}, c_{b2}, c_{b3}, c_{b4}$ | 12,25707814 | 0,45383519 | 0,84128534 | 0,00582191 | 0,06097942 |
| $c_{1b}, c_{11}, c_{12}, c_{13}, c_{14}$ | -0,44958276 | -0,31610971 | 0,76110175 | 0,00040195 | 0,00418876 |
| $c_{2b}, c_{21}, c_{22}, c_{23}, c_{24}$ | -1,47794571 | 0,86306804 | 0,60371593 | 0,00097653 | 0,01018521 |
| $c_{3b}, c_{31}, c_{32}, c_{33}, c_{34}$ | -0,92559941 | -0,08983701 | -0,19813900 | -1,80663161 | 3,02020703 |
| $c_{4b}, c_{41}, c_{42}, c_{43}, c_{44}$ | -4,65494620 | -0,45265677 | -0,99893282 | 2,39586683 | 8,59966897 |
| | | | | | $v(\chi^2)$ |
|  |  |  |  |  | **0,33** |
| **Ga-3-3** |  |  |  |  |  |
| $k_{be}$ | 0,01916959 | | | | |
| $k_{1b}, k_{2b}, k_{3b}, k_{4b}$ | 0,34516116 | 0,26298032 | 0,15272322 | 0,00021622 | |
| $k_{b1}, k_{b2}, k_{b3}, k_{b4}$ | 0,04445203 | 0,02453410 | 0,03317489 | 0,69473395 | |
| $\lambda_b, \lambda_1, \lambda_2, \lambda_3, \lambda_4$ | -0,86369475 | -0,32106156 | -0,24897878 | -0,14340459 | -0,00000580 |
| $c_{bb}, c_{b1}, c_{b2}, c_{b3}, c_{b4}$ | 11,01221836 | 0,40489350 | 0,33079422 | 0,19849340 | 0,00460051 |
| $c_{1b}, c_{11}, c_{12}, c_{13}, c_{14}$ | -0,94403811 | 0,74683138 | 0,15288116 | 0,04373307 | 0,00059249 |
| $c_{2b}, c_{21}, c_{22}, c_{23}, c_{24}$ | -0,44975594 | -0,17103110 | 0,57963170 | 0,04072613 | 0,00042920 |
| $c_{3b}, c_{31}, c_{32}, c_{33}, c_{34}$ | -0,51384499 | -0,07979346 | -0,11400965 | 0,70664873 | 0,00099937 |
| $c_{4b}, c_{41}, c_{42}, c_{43}, c_{44}$ | -8,86016462 | -0,87672540 | -0,92382864 | -0,96306777 | 15,18978642 |
| | | | | | $v(\chi^2)$ |
|  |  |  |  |  | **2,28** |
| **Ga-6-3** |  |  |  |  |  |
| $k_{be}$ | 0,01285646 | | | | |
| $k_{1b}, k_{2b}, k_{3b}, k_{4b}$ | 0,11015915 | 0,05620284 | 0,02992909 | 0,00019661 | |
| $k_{b1}, k_{b2}, k_{b3}, k_{b4}$ | 0,04085516 | 0,01903479 | 0,01169943 | 0,23716418 | |
| $\lambda_b, \lambda_1, \lambda_2, \lambda_3, \lambda_4$ | -0,34566051 | -0,09308429 | -0,05114738 | -0,02819543 | -0,00001009 |
| $c_{bb}, c_{b1}, c_{b2}, c_{b3}, c_{b4}$ | 13,28854644 | 0,84222053 | 0,32437035 | 0,12183425 | 0,01302843 |
| $c_{1b}, c_{11}, c_{12}, c_{13}, c_{14}$ | -2,30531841 | 2,01518848 | 0,22456880 | 0,06072879 | 0,00483235 |
| $c_{2b}, c_{21}, c_{22}, c_{23}, c_{24}$ | -0,87385720 | -0,43467623 | 1,22131747 | 0,08280270 | 0,00441326 |
| $c_{3b}, c_{31}, c_{32}, c_{33}, c_{34}$ | -0,49240722 | -0,15602042 | -0,17885270 | 0,82218575 | 0,00509459 |
| $c_{4b}, c_{41}, c_{42}, c_{43}, c_{44}$ | -9,12271070 | -2,15038792 | -1,50986982 | -1,03199790 | 16,56596634 |
| | | | | | $v(\chi^2)$ |
|  |  |  |  |  | **2,33** |

Table B 1: Coefficients of the optimized 5-compartment model.
|  |  |  |  |  |  |
| --- | --- | --- | --- | --- | --- |
| $k_{be}$ | 0,03711453 | | | | |
| $k_{1b}, k_{2b}, k_{3b}, k_{4b}$ | 0,30251036 | 0,11734788 | 0,03929789 | 0,00016964 | |
| $k_{b1}, k_{b2}, k_{b3}, k_{b4}$ | 0,15308706 | 0,22169987 | 0,09084287 | 0,30280958 | |
| $\lambda_b, \lambda_1, \lambda_2, \lambda_3, \lambda_4$ | -0,91749277 | -0,24357421 | -0,07568756 | -0,02810666 | -0,00001848 |
| $c_{bb}, c_{b1}, c_{b2}, c_{b3}, c_{b4}$ | 14,46366478 | 1,05210959 | 0,86157135 | 0,50424105 | 0,00941323 |
| $c_{1b}, c_{11}, c_{12}, c_{13}, c_{14}$ | -3,60042814 | 2,73286213 | 0,58149108 | 0,28131102 | 0,00476391 |
| $c_{2b}, c_{21}, c_{22}, c_{23}, c_{24}$ | -4,00751493 | -1,84789149 | 4,58494532 | 1,25267431 | 0,01778678 |
| $c_{3b}, c_{31}, c_{32}, c_{33}, c_{34}$ | -1,49616094 | -0,46787930 | -2,15081966 | 4,09308959 | 0,02177031 |
| $c_{4b}, c_{41}, c_{42}, c_{43}, c_{44}$ | -4,77447487 | -1,30888613 | -3,45470371 | -5,46547147 | 18,85653618 |
| | | | | | $v(\chi^2)$ |
|  |  |  |  |  | **0,37** |

**Fig. B 1:**
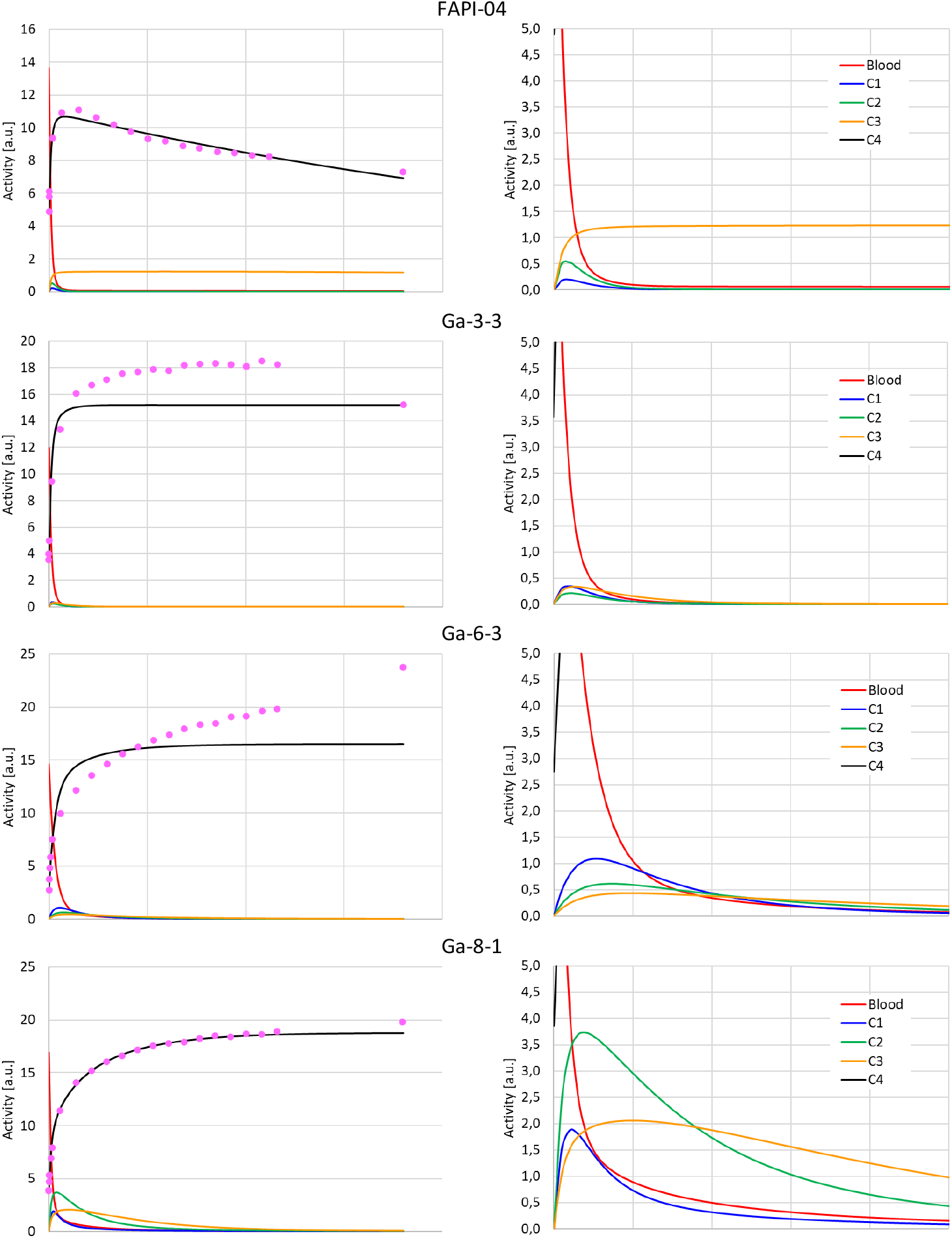
Blood and tumor TACs of the optimized 5-compartment model based on the data measured by Wang et al. [10]

For the second and third model with matrix 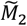 and 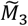 similar problems were observed. These findings suggest that standard 5-compartment models are not suitable to reflect the particular pharmacokinetic processes that underlie the experimental data of Wang et al.

